# Decoding Fibrosis: Transcriptomic and Clinical Insights via AI-Derived Collagen Deposition Phenotypes in MASLD

**DOI:** 10.1101/2025.08.29.25334719

**Authors:** Marta Wojciechowska, Mira Thing, Yang Hu, Gianluca Mazzoni, Lea Mørch Harder, Mikkel Parsberg Werge, Nina Kimer, Vivek Das, Jaime Moreno Martinez, Cesar Prada-Medina, Mogens Vyberg, Robert Goldin, Reza Serizawa, Jeremy Tomlinson, Elisabeth Douglas Galsgaard, Dan J Woodcock, Henning Hvid, Dominik Reinhard Pfister, Vanessa Isabell Jurtz, Lise Lotte Gluud, Jens Rittscher

## Abstract

Histological assessment is foundational to multi-omics studies of liver disease, yet conventional fibrosis staging lacks resolution, and quantitative metrics like collagen proportionate area (CPA) fail to capture tissue architecture. While recent AI-driven approaches offer improved precision, they are proprietary and not accessible to academic research. Here, we present a novel, interpretable AI-based framework for characterising liver fibrosis from picrosirius red (PSR)-stained slides. By identifying distinct data-driven collagen deposition phenotypes (CDPs) which capture distinct morphologies, our method substantially improves the sensitivity and specificity of downstream transcriptomic and proteomic analyses compared to CPA and traditional fibrosis scores. Pathway analysis reveals that CDPs 4 and 5 are associated with active extracellular matrix remodelling, while phenotype correlates highlight links to liver functional status. Importantly, we demonstrate that selected CDPs can predict clinical outcomes with similar accuracy to established fibrosis metrics. All models and tools are made freely available to support transparent and reproducible multi-omics pathology research.

**Highlights:**

- We present a set of data-driven collagen deposition phenotypes for analysing PSR-stained liver biopsies, offering a spatially informed alternative to conventional fibrosis staging and CPA available as open-source code.
- The identified collagen deposition phenotypes enhance transcriptomic and proteomic signal detection, revealing active ECM remodelling and distinct functional tissue states.
- Selected phenotypes predict clinical outcomes with performance comparable to fibrosis stage and CPA, highlighting their potential as candidate quantitative indicators of fibrosis severity.

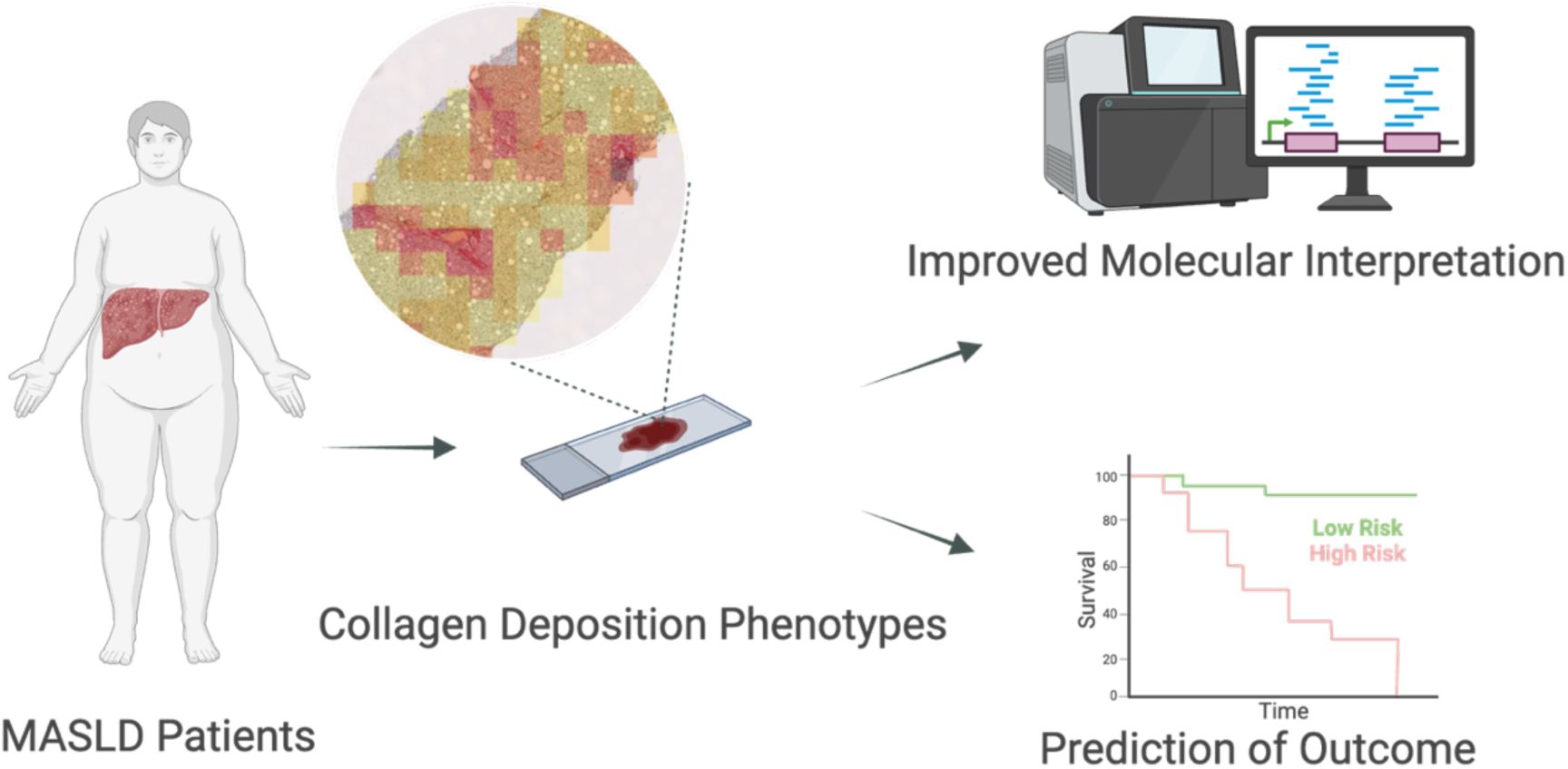

## Introduction

Metabolic dysfunction-associated steatotic liver disease (MASLD) is the most common chronic liver disease worldwide, contributing to liver-related, cardiovascular, and all-cause mortality (1–3). While most cases are mild, some progress to severe complications such as cirrhosis and hepatocellular carcinoma (HCC). Identifying high-risk patients requires balancing detection with avoiding unnecessary tests and overtreatment (4).

Despite advances in non-invasive tests (NITs) like circulating biomarkers and imaging, they remain insufficient for definitive diagnosis or staging (5,6). While NITs aid screening and monitoring (7), histology remains the gold standard for directly evaluating steatosis, inflammation, ballooning, and fibrosis (7). NIT-based fibrosis scores can be affected by age and comorbidities, potentially leading to misclassification of disease severity (6). Since histological fibrosis stage is a strong predictor of liver-related outcomes, histological evaluation remains important for accurate disease stratification and determining eligibility for clinical trials (9,10).

### Rationale for AI-driven phenotyping of fibrosis morphology

Traditional histological evaluation relies on subjective interpretation by pathologists, leading to interobserver variability and inconsistencies in categorical grading (11,12). Integrating AI into histology offers a promising route to improved diagnostic accuracy (12). Grading systems like the NASH CRN (13) use rigid categories that fail to capture fibrosis as a continuum. Fibrogenesis is a dynamic process and conventional staging (F0–F4) may not fully capture collagen deposition patterns. Categorical staging lacks the granularity to track fibrosis progression or regression (12,14). These limitations impact both clinical practice and research into MASLD pathogenesis.

Quantitative histology enables continuous fibrosis assessment using metrics like collagen proportionate area (CPA) (15,16). CPA predicts outcomes in MASLD (17) and other liver diseases (18,19) but does not capture spatial collagen organisation or features like chicken-wire fibrosis, bridges, or septa, key elements in traditional scoring (20). Recent studies have proposed alternative AI-based fibrosis models which partly address these limitations (21,22).

AI can detect complex, prognostically relevant fibrosis patterns, enabling a more nuanced assessment of disease burden. This may enhance fibrosis evaluation and aid in identifying histology-based endpoints, biomarkers, and predictive models.

### Existing liver fibrosis algorithms

Digital pathology methods for liver fibrosis fall into two categories: “AI pathologist” models, which automate scoring (e.g. NAS score) (23–27), and quantitative approaches producing continuous metrics like CPA (15). While useful for automation, AI-pathologist models replicate existing systems without adding new insights. In contrast, CPA is a spatially blind metric which on its own does not encode important morphological features of fibrosis. Recent tools attempt to combine qualitative and quantitative aspects of fibrosis assessment (21,22,28–31). These tools often extract fibre features (e.g. length, thickness) and compress them into composite scores (22,29,30). The drawback of such abstract metrics is their lack of interpretability. Another approach uses deep learning to extract fibrosis features and classify tiles post hoc, e.g. by NASH CRN score (21). Commercial platforms often offer a combination of approaches and do not list implementation details transparently (21,22,29,30), which means they cannot be reproduced.

### MASLD omics studies

Large-scale studies have examined the molecular landscape of liver fibrosis via transcriptomics and proteomics (32–37). Notably, Govaere et al. provided key insights into gene expression across the fibrosis spectrum (33,34). However, these studies did not incorporate spatially resolved histological features such as subtypes of collagen morphology. Our work builds on these efforts by integrating deep-learning-based spatial characterisation of fibrosis, offering a more nuanced and biologically grounded histological framework for omics integration.

Blood proteomics is particularly attractive for developing non-invasive liver tests due to the predominantly liver-derived nature of blood proteins (38). We explore tissue transcriptomic and proteomic correlates of collagen deposition phenotypes (CDPs) to examine fibrosis patterns across disease stages.

### Hypothesis and study objectives

The objective of this study is three-fold. Firstly, we provide a novel, open, AI-based method for quantification of fibrosis in liver histology slides in the form of CDPs. Secondly, we utilise the notion of these phenotypes in a study of transcriptomic and proteomic features of MASLD. Finally, we demonstrate that CDPs can predict patient outcome in our study cohort of 187 MASLD patients.

## Methods

### Participants and study design

We included 187 MASLD patients (F0: 35, F1: 53, F2: 50, F3: 24, F4: 25) and 15 healthy volunteers. MASLD patients had a mean age of 51.3 years (±14.2) and BMI of 33.3 kg/m² (±6.32); healthy volunteers were 31.5 (±17.0) with BMI 25.0 kg/m² (±3.97). Baseline characteristics are summarized in Table 1.

**Table 1.** CoCoMASLD cohort baseline characteristics.

|  | MASLD<br>N = 187 | Healthy Volunteer<br>N = 15 |
| --- | --- | --- |
| Sex, Male, n (%) | 105 (56.1%) | 9 (60.0 %) |
| Age, years, Median [IQR] | 54 [41.0, 62.0] | 25.0 [21.0, 30.50] |
| Ethnicity, Caucasian, n (%) | 148 (79.1%) | 14 (93.3%) |
| BMI, kg/m<sup>2</sup>, mean (SD) | 33.3 (6.32) | 25 (3.97) |
| Dyslipidemia, n (%) | 112 (59.9%) | 0 (0%) |
| Type 2 diabetes, n (%) | 83 (44.4%) | 0 (0%) |
| Hypertension, n (%) | 89 (47.6 %) | 0 (0%) |
| Cardiovascular disease, n (%) | 23 (12.3%) | 0 (0%) |
| LSM baseline, kPa, Median [IQR] | 9.80 [6.58, 14.43] | 4.30 [3.65, 4.80] |
| Fib4, Median [IQR] | 1.29 [0.78 – 1.83] | 0.65 [0.44, 0.84] |
| ALT, U/L, mean (SD) | 81.7 (65.3) | 19.6 (6.33) |
| AST, U/L, mean (SD) | 55.0 (40.0) | 22.3 (5.74) |
| Platelets, 10<sup>9</sup>/L, mean (SD) | 238 (71.6) | 242 (55.2) |
| INR, mean (SD) | 1.01 (0.14) | 1.03 (0.09) |
| Albumin, g/L, mean (SD) | 39.3 (3.62) | 40.5 (2.81) |
| Bilirubin, μmol/L, mean (SD) | 10.9 (6.39) | 10.8 (8.02) |
| Fibrosis Score, n (%) |  |  |
| F0 | 35 (18.7%) | 15 (100%) |
| F1 | 53 (28.3%) | 0 (0%) |
| F2 | 50 (26.7%) | 0 (0%) |
| F3 | 24 (12.8%) | 0 (0%) |
| F4 | 25 (13.4%) | 0 (0%) |
| Patients with an LRE | 14 |  |
| Liver related death | 7 |  |
| HCC | 0 |  |
| Liver transplantation | 0 |  |
| Hepatic encephalopathy | 5 |  |
| Hepatorenal syndrome | 3 |  |
| Variceal hemorrhage | 3 |  |
| Varices on endoscopy | 10 (1 not having other LREs) |  |
| Ascites | 8 |  |
| Jaundice | 9 |  |

Participants were enrolled in the prospective Copenhagen Cohort of Metabolic Dysfunction Associated Steatotic Liver Disease (CoCoMASLD), designed to evaluate clinical predictors and biomarkers. The study was approved by the Regional Committee on Health Research Ethics (H-17029039) and conducted in accordance with the Declaration of Helsinki and all participants provided their informed consent. Patients were recruited from the outpatient department at the Gastro Unit, Copenhagen University Hospital Hvidovre, Denmark. Healthy volunteers were recruited through advertisements. All participants underwent clinical assessment, blood tests, and liver biopsy. Healthy volunteers all had transjugular liver biopsies (TJLB), while 33 patients underwent TJLB, and 154 patients had percutaneous liver biopsies. Tissue sections were stained for collagen with PicroSirius Red and scanned at 40x (Hamamatsu C13210). The PSR-stained histology slides were scored for fibrosis by two pathologists according to the Kleiner-Brunt system.

At baseline, all participants underwent liver stiffness measurement (LSM) using vibration-controlled transient elastography (FibroScan®), along with blood sample collection. Additionally, patients with MASLD had LSM and blood samples taken at yearly follow-up visits.

### AI-Derived Collagen Phenotypes

To investigate the spatial architecture of liver fibrosis in the CoCoMASLD cohort, we developed a deep learning method to identify distinct patterns of collagen deposition from PSR-stained liver biopsies. The workflow consists of three steps corresponding to the panels in Figure 1: collagen segmentation and tile extraction (A), phenotyping of collagen deposition (B), and slide-level spatial classification (C).

**Figure 1.**
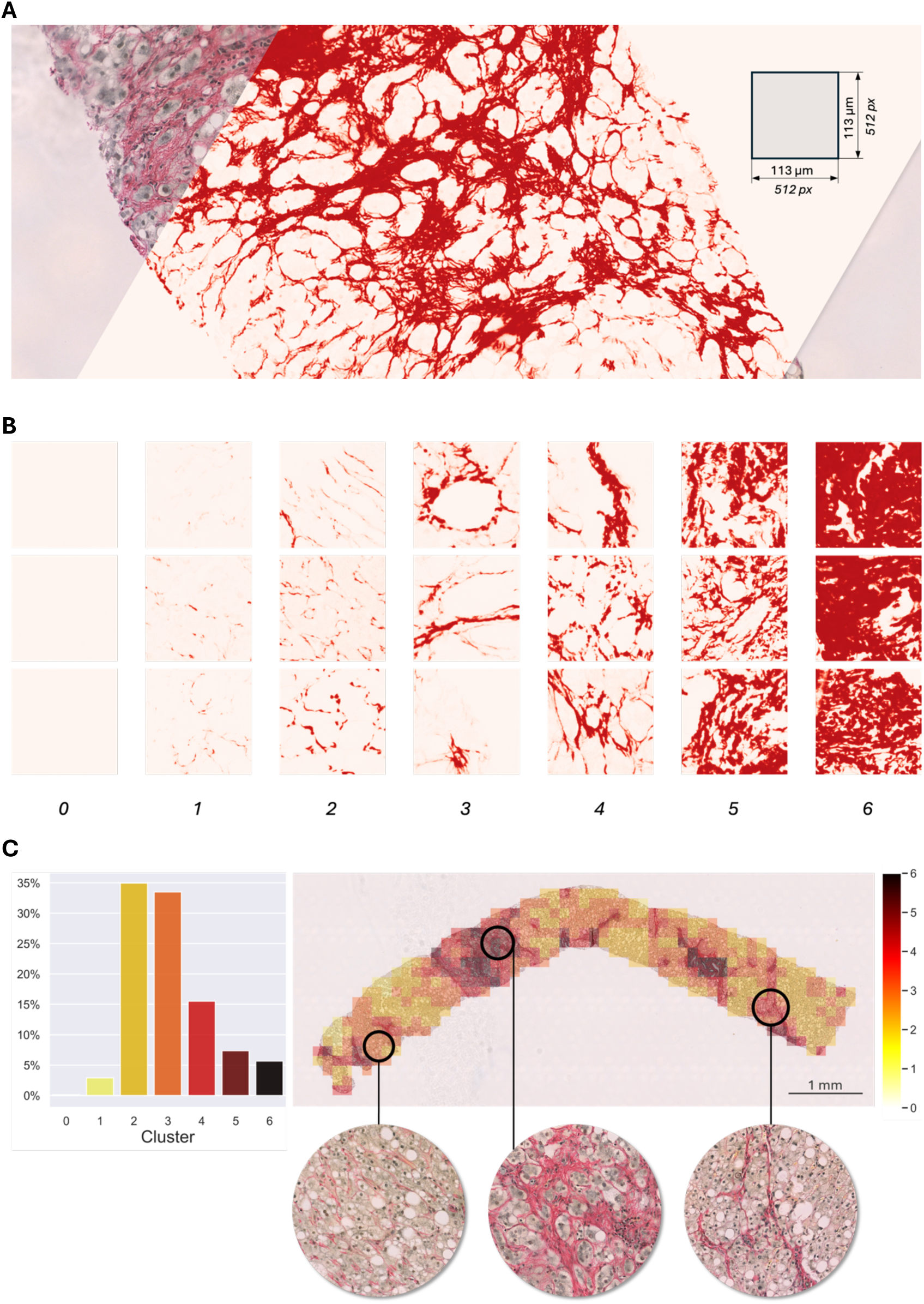
A digital pipeline for spatial collagen phenotyping in liver tissue. **A)** Deep learning-based collagen segmentation on PSR-stained slides enables robust fibre detection across the biopsy. **B)** AI-feature-based collagen phenotyping captures distinct morphologies, from sparse fibrils (clusters 0–2) to dense aggregates (clusters 4–6). **C)** These collagen subtypes are spatially mapped across the tissue, allowing quantification of their relative abundance and localisation patterns.

### Collagen segmentation and tile extraction

A convolutional neural network (U-Net) (39) was used to segment collagen from PSR-stained slides (Figure 1A). U-Net and U-Net derived segmentation models are not only widely available they are now accepted as a reference architecture for biomedical image segmentation (40). This image-filtering approach is well-suited to handle stain variability across samples and provides a robust and reproducible way to compute collagen proportionate area (CPA) (41). Segmentation ensured downstream analyses were based solely on PSR, unaffected by confounding features like nuclei or fat vesicles. Tiles (113 × 133 µm) were extracted from all tissue-positive regions, regardless of collagen content.

### Deep clustering of collagen deposition phenotypes

The tile size was chosen to match the scale of anatomical and pathological features relevant to observing the progression of fibrogenesis, such as portal tract edges, central veins, and fibrotic bridges. This resolution captures local ECM features while preserving enough context for meaningful morphology classification. Tiles were embedded into a high-dimensional space using ResNet18 (42) pretrained on ImageNet (43). Feature vectors were clustered via k-means into collagen morphology subtypes.

### Slide-level spatial classification

The trained model assigned each tile to a collagen deposition phenotype, generating spatial maps of ECM distribution in all CoCoMASLD slides. For every case, the relative abundance of each phenotype was computed (Figure 1C). Case-level phenotypic profiles were encoded as vectors for downstream multi-omic and outcome analyses. Model and implementation details are in the Supplementary Material.

### Comparison of Fibrosis Metrics and Their Gene Expression Profiles

RNA-seq was performed on FFPE liver biopsies using the TruSeq RNA Exome kit. Serum protein levels were measured using the SomaScan v4.1 (7k) platform. A custom pipeline (see Supplementary Material) was used for RNA and protein differential expression analysis.

To compare the utility of distinct fibrosis metrics in identifying gene expression changes, we applied a consistent statistical and effect size threshold across all differential gene expression (DGE) analyses. Transcripts with adjusted p < 0.05 and absolute fold-change > 1 (i.e., more than twofold difference in expression) were retained. Effect sizes were presented on a linear, non-log-transformed scale, ensuring comparability across metrics derived from categorical contrasts (e.g., no fibrosis vs. advanced fibrosis (F3-F4)) and continuous variables (e.g., CPA, CDP proportions). While the threshold does not carry the same biological interpretation across all models, it serves as a practical benchmark for identifying robust transcriptional shifts.

We used Venn diagrams to show gene-level overlap between C4–C6, CPA, and fibrosis score. The diagrams were generated to illustrate the shared and unique genes between these fibrosis metrics, providing insight into their complementarity. Pearson’s correlation was used to assess the degree of concordance between the metrics, and statistical significance was determined by permutation testing.

### Pathway Enrichment Analysis

We performed pathway overrepresentation analysis (ORA) on gene expression data from CDPs C4–C6 to explore fibrosis-related pathways. The analysis focused on identifying pathways whose enrichment correlated with an increased proportion of specific phenotypes, relative to the entire annotated gene set. We used Reactome’24 to explore enrichment across a range of biological processes, such as extracellular matrix (ECM) organisation, immune response, and metabolic pathways (41).

We applied a false discovery rate (FDR) threshold of < 0.05 to determine statistical significance for pathway enrichment. Pathways below this threshold were deemed significantly enriched. For comparison, we also performed the same analysis on CPA and fibrosis score to assess how these traditional metrics align with the pathway signals identified from CDPs.

To identify key fibrosis-associated molecules, we compiled tables of the most up- and downregulated genes and proteins across metrics. Significant genes and proteins (adjusted p < 0.05) were ranked by effect size. For each metric, the top 10 molecules are presented.

### Clinical outcomes

All participants were included in the baseline evaluation of correlations between AI-derived histology metrics, CRN fibrosis stages, and LSM. Only MASLD patients were included in the analyses of disease progression. All statistical analyses were conducted using R v4.3.0. Univariable and multivariable competing risk Cox proportional hazards models, adjusted for age and sex, were employed in the time-to-event analyses. Disease progression was defined as the occurrence of a liver-related event (LRE) after inclusion. LREs included liver-related death, liver transplantation, hepatocellular carcinoma, hepatic encephalopathy, hepatorenal syndrome, jaundice, ascites, variceal haemorrhage, and varices detected on endoscopy. Receiver operating characteristic (ROC) curves were generated to determine Youden’s index thresholds, used in Kaplan-Meier plots. Age and sex adjusted logistic regression models were used to predict odds ratios (OR) of a clinically significant increase in liver stiffness (LSM), defined as an increase of ≥5 kPa and ≥20% from the baseline LSM value. For further statistical methods see supplementary material.

## Results

Our tile feature phenotyping pipeline identified seven distinct collagen morphology sub-types (C0–C6). These ranged from liver tissue tiles with no identifiable collagen, through regions with intact, non-fibrotic ECM, to progressively denser fibrotic patterns including bridging fibrosis and thick collagen aggregates (Figure 1B). While individual phenotypes do not directly map to specific anatomical structures such as portal tracts or central veins, the extreme ends of the spectrum, those with either little identifiable collagen or with dense fibrotic aggregates, are more readily interpretable in biological terms.

Of the 202 participants enrolled in the study, 4 cases were excluded due to slide staining issues, 6 had no available PSR-stained slides, 2 digitised slides were corrupted, and 2 additional cases failed image processing. In total, CDP computation was successfully performed on 188 cases.

We investigated the transcriptomic and proteomic signatures associated with fibrosis progression across different fibrosis metrics, including CDPs (C0–C6), collagen proportionate area (CPA), and fibrosis score. Our analysis revealed that CDPs (C4–C6) capture the most substantial molecular signals across transcriptomic data, providing insights into key fibrotic processes (Figure 2A). In contrast, C0–C3, which represent early or sparse collagen deposition, show minimal effect sizes, indicating minimal transcriptional deviation compared to average liver tissue in the CoCoMASLD cohort. This suggests that these areas do not exhibit the robust transcriptional shifts typically associated with fibrosis progression.

**Figure 2.**
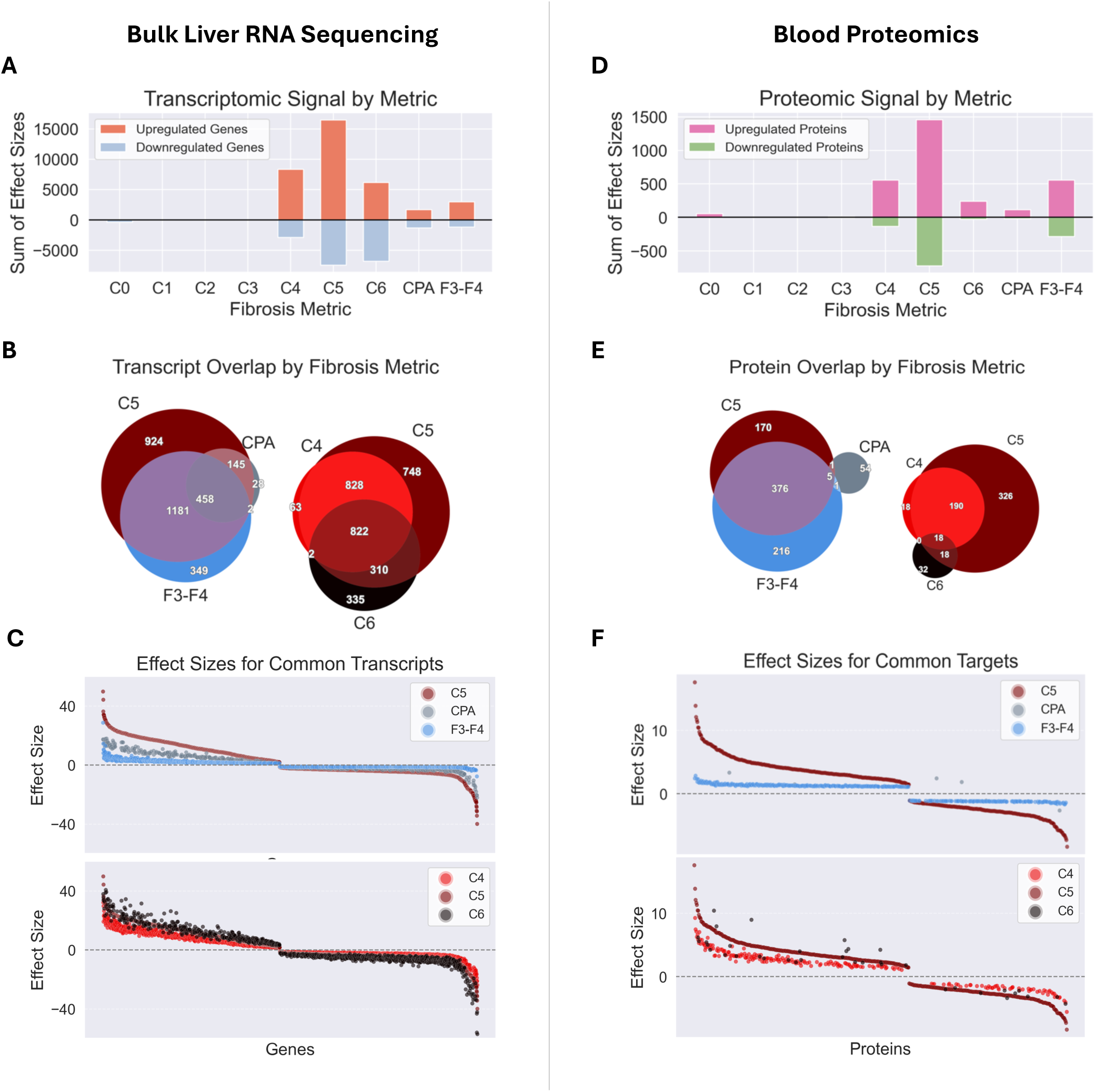
CDP 5 captures the strongest molecular signals across fibrosis metrics. **A)** CDP C5 captures the largest cumulative transcriptomic signal across metrics. **B)** Transcript-level overlap reveals shared molecular signatures. **C)** Shared transcripts exhibit concordant effect sizes, with CDP C5 consistently producing the strongest associations. **D–F)** Proteomic patterns mirror transcriptomic results. **D)** CDP C5 shows the strongest cumulative protein-level effect size. **E)** Protein overlap indicates stronger alignment between CDP C5 and F3–F4 than between CPA and F3–F4. **F)** Among shared proteins, CDP C5 yields the largest effect sizes.

Further investigation into gene overlap (Figure 2B) revealed that C5, fibrosis score, and CPA shared a substantial set of differentially expressed genes, suggesting common biological pathways across these metrics. Similarly, C4, C5, and C6 share many genes between each other, yet each also exhibits distinct molecular signatures, suggesting that CDPs capture different aspects of fibrosis progression.

The concordance of effect sizes (Figure 2C) further reinforced these findings, with C5 showing the most pronounced molecular shifts for each transcript, compared to the same transcripts in CPA and F3–F4. This demonstrates that C5 yields a stronger molecular signal across fibrosis metrics for the same set of differentially expressed genes.

To evaluate the fidelity of CDPs as fibrosis descriptors, we benchmarked CDPs C4, C5, and C6, along with collagen proportionate area (CPA), against the transcriptomic signature of advanced fibrosis (F3–F4) (Figure 2C). All four metrics showed strong and statistically significant positive correlations with the F3–F4 DGE profile. Phenotype C5 demonstrated the highest concordance with advanced fibrosis (Pearson r = 0.948, p<0.001). C4 and C6 followed closely, and all outperformed or matched CPA (r=0.878, p<0.001). These results support the utility of the morphology-based CDPs, particularly C5, as biologically robust markers of fibrotic severity.

We next explored the proteomic data to assess the molecular signals captured by the fibrosis metrics. Like the transcriptomic data, C5 exhibits the largest cumulative protein-level effect size (Figure 2D), reinforcing its role in capturing significant biological processes associated with fibrosis. Further analysis of protein-level overlap (Figure 2E) revealed a stark contrast in the protein sets: C5 shares 376 proteins with F3–F4, while CPA shares only 6 proteins with each. Unlike CDPs C4–C6, CPA shows limited overlap with fibrosis-associated transcripts and proteins, suggesting it lacks sensitivity to active fibrogenic processes.

Further investigation into protein-level overlap between C4, C5, and C6 (Figure 2B) revealed that, similar to the transcriptomic data, these phenotypes share many of the same protein targets. Interestingly, C4 is almost entirely contained within C5, with 208 shared proteins and only 18 proteins exclusive to C4. This suggests that C5 captures a broader and more comprehensive fibrosis-related protein signal, while C4 primarily reflects a subset of the proteins found in C5. In contrast, C6 shows more distinct molecular associations, further highlighting the unique CDP signatures.

### Phenotype specific molecular activity

Pathway overrepresentation analysis has shown that C4 and C5 exhibit enrichment across all the annotated extracellular matrix (ECM) remodelling pathways, highlighting that these phenotypes represent regions where fibrosis-related processes occur (Figure 3A). While C5 shows the strongest and most widespread enrichment, C4 follows a similar signature but with less pronounced activation, corresponding to histological evidence suggesting that C4 reflects a transitional stage between early and advanced fibrosis.

**Figure 3.**
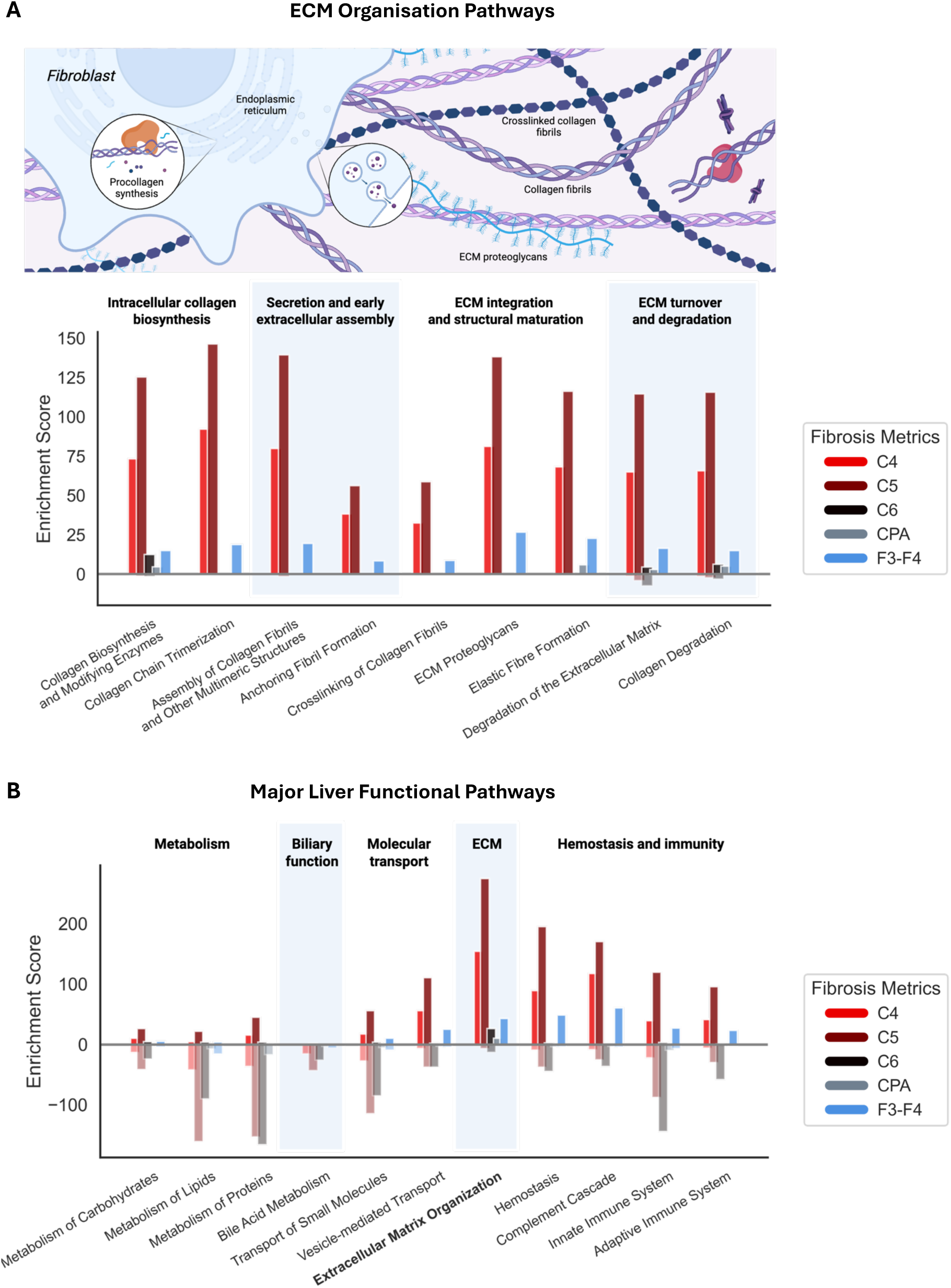
CDPs uncover distinct biological programmes in fibrosis. A) CDP C5 shows widespread enrichment in ECM remodelling pathways. CDP C4 exhibits similar but weaker activation. CDP C6 reveals a distinct ECM signature, suggesting a divergent tissue state. B) Transcriptomic pathway enrichment across liver systems. CDPs C4–C6 show broader activation across metabolic, immune, and transport pathways compared to CPA or histological staging. All pathways shown meet FDR < 0.05.

In contrast, C6 shows very little enrichment in any of the shown ECM remodelling pathways, suggesting that little to no active fibrogenesis occurs in these dense scaring regions, which lacks hepatocytes. This supports the understanding that C6 represents a late-stage fibrotic tissue, where ECM deposition has already occurred, and the active remodelling processes are minimal.

Further analysis of broader transcriptomic remodelling (Figure 3B) across liver systems revealed that CDPs (C4–C6) exhibited stronger and more diverse pathway activation compared to CPA or fibrosis score. Notably, C4–C6 showed significant enrichment across metabolic, immune, and transport processes, which are known to play a key role in metabolic dysfunction-associated steatohepatitis (MASH) progression. In particular, C6 shows downregulation across key liver functional pathways, particularly in metabolism, molecular transport, and immune response. This finding is consistent with impaired liver function observed in cirrhosis, suggesting that C6 represents a non-progressive, non-functional tissue, obstructing normal liver function.

These findings underscore the sensitivity of CDPs in capturing the complex molecular signals of fibrosis and highlight their utility as discovery tools for identifying new fibrosis-related biological pathways.

### Differentially expressed molecules

Many of the upregulated transcripts identified by CDPs (Figure 4) confirm earlier findings. STMN2 (34), *KRT23* (*44*), *FGF7* (45), *SMOC2* (*46*), *FAP* (*47*) are all expected in the liver fibrosis context. Similarly, MFAP4 (48) and GDF15 (16) have been previously identified as potential blood biomarkers for liver cirrhosis. Unlike CPA, which does not show significant enrichment of *KRT23*, *FGF7*, *FAP*, MFAP4, or GDF15, the CDPs (C4, C5) and advanced fibrosis stage (F3–F4) highlight these molecules, underscoring the added specificity of our method.

**Figure 4.**
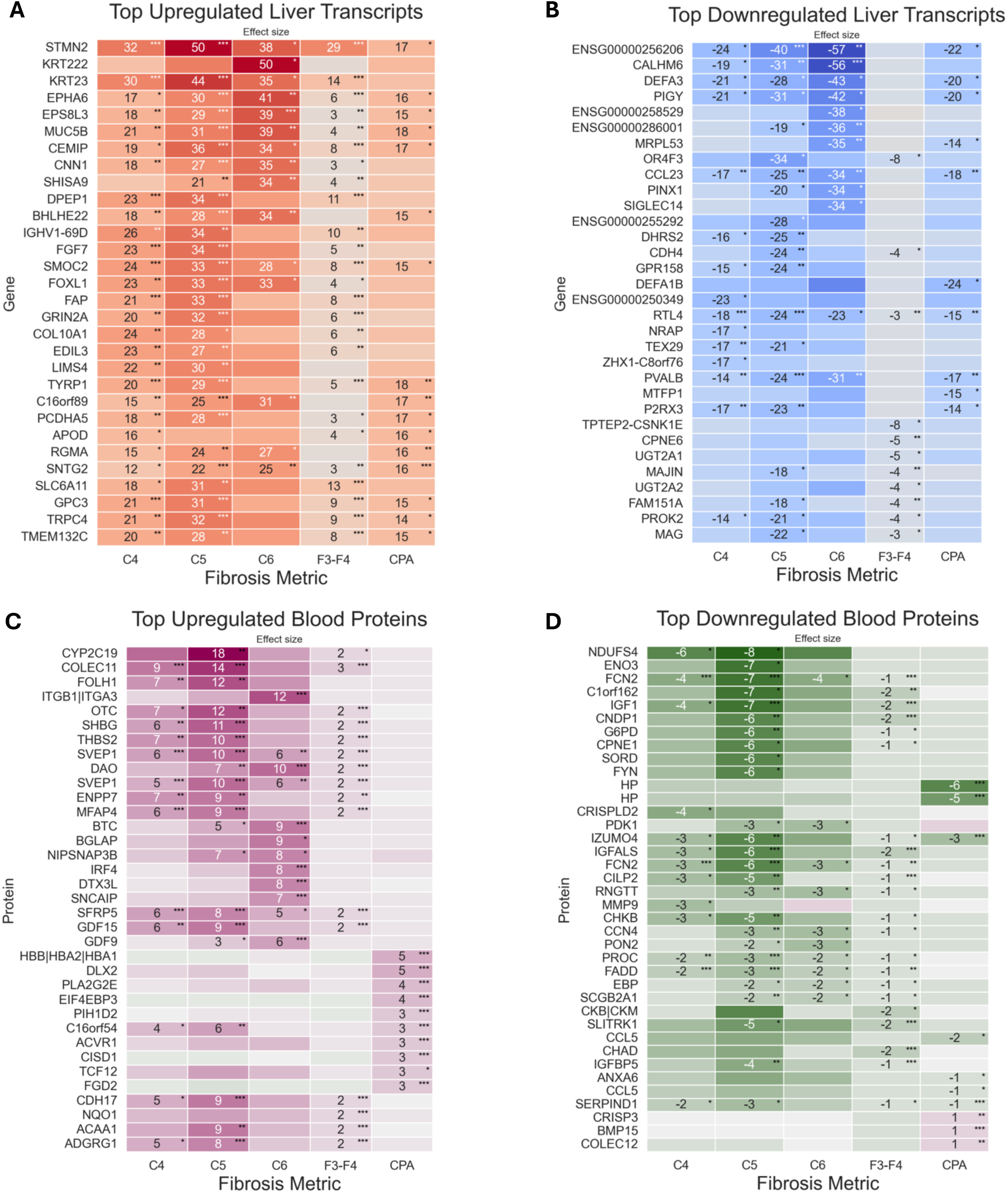
Differentially expressed molecules across fibrosis metrics. **A)** Top upregulated transcripts show consistent enrichment in ECM remodelling genes. CDPs reflect refined collagen deposition subtypes not captured by CPA, offering a more detailed molecular characterisation of fibrosis patterns. **B)** Top upregulated proteins by fibrosis metric. CDPs C4 and C5 show a strong correlation with fibrosis score, while CPA deviates from this trend. This suggests that CDPs capture fibrosis-associated molecular changes more specifically than CPA.

Using the top 100 upregulated transcripts from C5 as a reference, we computed Jaccard indices to assess gene-level concordance with other fibrosis metrics. Phenotype C4 showed the strongest overlap (Jaccard index = 0.64), sharing nearly two-thirds of C5’s top upregulated genes. F3–F4 also demonstrated substantial concordance (0.46), while CPA (0.19) and C6 (0.18) showed only limited overlap. These findings suggest that C5 and C4 capture a closely related fibrotic transcriptional programme.

Applying the same approach to plasma proteomics, we compared the top 100 upregulated circulating proteins from C5 against those from other metrics. C4 and F3–F4 again showed strong concordance with C5 (Jaccard indices = 0.56 and 0.53, respectively), consistent with transcriptomic findings. In contrast, CPA (0.006) showed almost no overlap with C5’s plasma protein signature, suggesting that this area-based metric may capture architectural features that do not strongly engage systemic molecular responses. C6 (0.07) also showed minimal overlap, consistent with its histological interpretation as dense scar tissue characteristic of late-stage fibrosis, in which active transcriptional and proteomic changes may have largely subsided. Full results of the differential expression analyses for transcriptomic and proteomic datasets are provided in the Supplementary Data 1.

### Comparison with CPA

Our analysis revealed a gradual shift in collagen architecture with disease progression, from low-density fibrillar patterns (CDPs 0–2) in early-stage fibrosis to dense, aggregated patterns (CDPs 4–6) in advanced disease (Fig. 5A). Notably, collagen proportionate area (CPA), a commonly used digital fibrosis marker, was disproportionately influenced by low-density collagen in healthy volunteers and in early disease stages (Fig. 5B).

**Figure 5.**
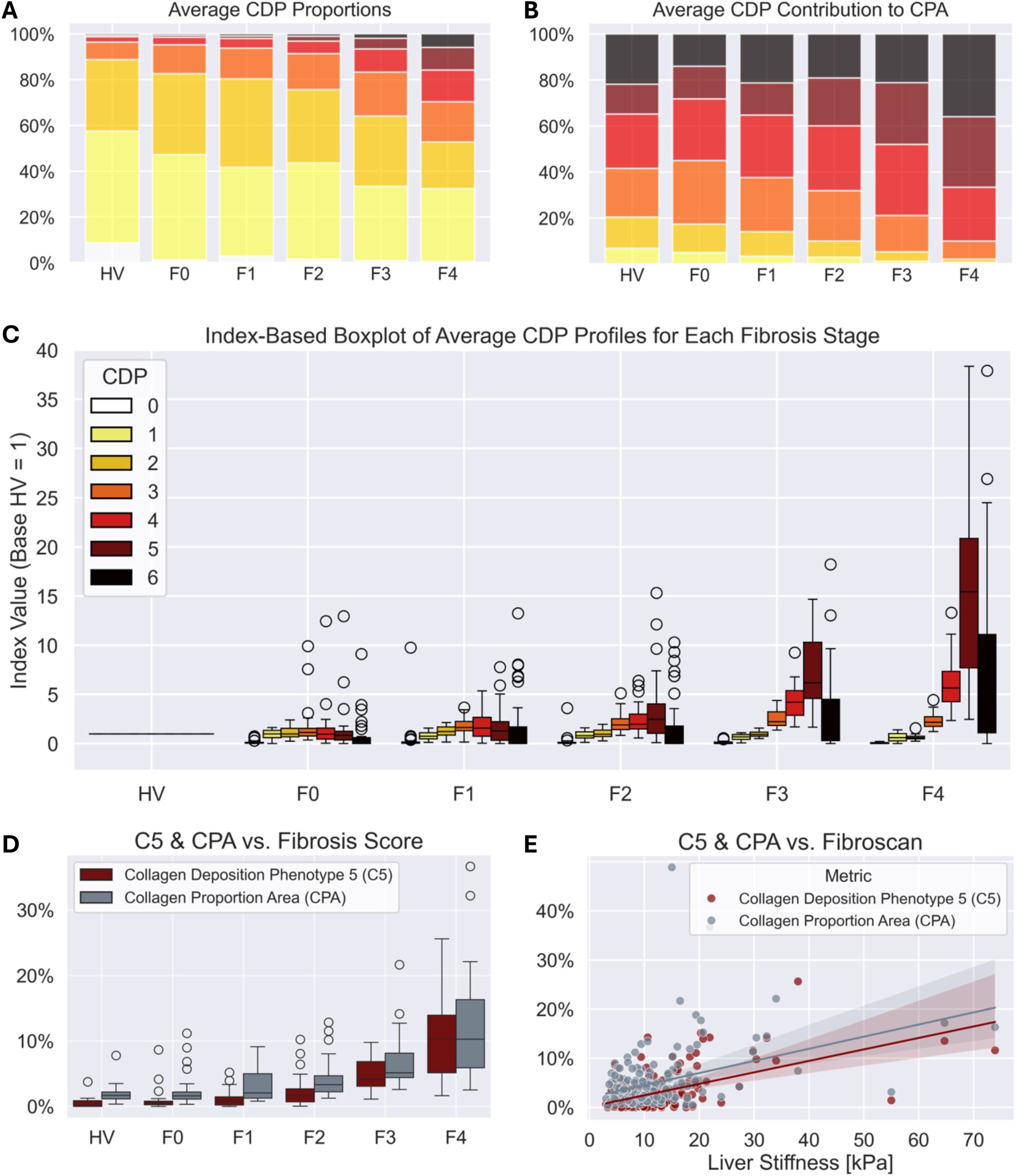
Digital quantification of CDPs reveals stage-specific fibrosis patterns. **A)** Fibrosis progression is marked by a gradual shift from low-to high-collagen clusters. **B)** CPA is disproportionately influenced by low-density collagen in early fibrosis, potentially inflating scores. **C)** CDP C5 shows the strongest stage-dependent increase among collagen morphology clusters. **D)** CDP C5 correlates more strongly with fibrosis stage than CPA (C5: r = 0.67; CPA: r = 0.42, p<0.001). **E)** CDP C5 correlates more strongly with liver stiffness than CPA (C5: r = 0.57; CPA: r = 0.39, p<0.001).

Because CPA aggregates all collagen regardless of its structural organization, it captures contributions from low-density fibrillar patterns that are prevalent even in healthy or minimally fibrotic tissue. As a result, CPA values can appear elevated in early-stage disease, despite the absence of advanced fibrotic remodelling (15). This compromises its specificity for capturing structural collagen remodelling associated with fibrosis progression and may lead to inflated estimates of disease burden or misinterpretation of treatment effects in both clinical and research settings.

Amongst all phenotypes, CDP 5 (C5) exhibited the strongest stage-dependent increase, making it a robust marker of stage-associated collagen remodelling (Fig. 5C). Consistent with this, C5 correlated more strongly than CPA with both histological fibrosis stage (r = 0.67*** vs. 0.42***) and liver stiffness measurements (r = 0.57*** vs. 0.39***; Fig. 2D–E). Compared to CPA, C5 provides a more specific readout of fibrosis progression and is more sensitive to advanced-stage collagen remodelling.

### Prediction of clinical outcome

In total, 14 (7.5%) MASLD patients experienced a liver related event (LRE) during a median follow-up of 4.79 years (IQR 4.09-5.74), corresponding to 940.69 patient-years. During follow-up, 13 (6.95%) MASLD patients died, of whom 7 (53.85%) had liver-related deaths. Among the 14 patients with a liver-related event, 5 developed hepatic encephalopathy, 3 had hepatorenal syndrome, 3 experienced variceal haemorrhage, 8 had ascites, 9 developed jaundices, and 10 had varices on endoscopy. Only one patient had varices on endoscopy as their sole liver-related event.

### Phenotype-specific hazard ratio of liver related events in MASLD

An increase in CDPs C4, C5 and C6 was associated with a higher hazard of LREs during follow-up. Since the differences between models were minor, results from the multivariable competing risk model are presented, while findings from the univariable model are shown in Supplementary Table 1. Each 1% increase in C4 was associated with a hazard ratio (HR) of 1.12 (95% CI: 1.05–1.20, *p* = 0.0005), in C5 with an HR of 1.21 (95% CI: 1.11–1.32, *p* < 0.0001), and in C6 with an HR of 1.13 (95% CI: 1.03–1.24, *p* = 0.010). In contrast, C1 was associated with a slightly reduced hazard of LRE (HR 0.96, 95% CI: 0.93–0.99, *p* = 0.022). No significant associations were observed for C0, C2, or C3 (Supplementary Table 1). Both the CPA (HR of 1.08 95% CI: 1.02–1.15, *p* = 0.014) and the CRN fibrosis score (HR 2.09, 95% CI: 1.26–3.46, *p* = 0.004) were also significantly associated with an increased hazard of LRE during follow-up. Results from competing risk cox regression analyses of dichotomized variables are shown in Table 2, and corresponding Kaplan-Meier plots are shown in Figure 6.

**Figure 6:**
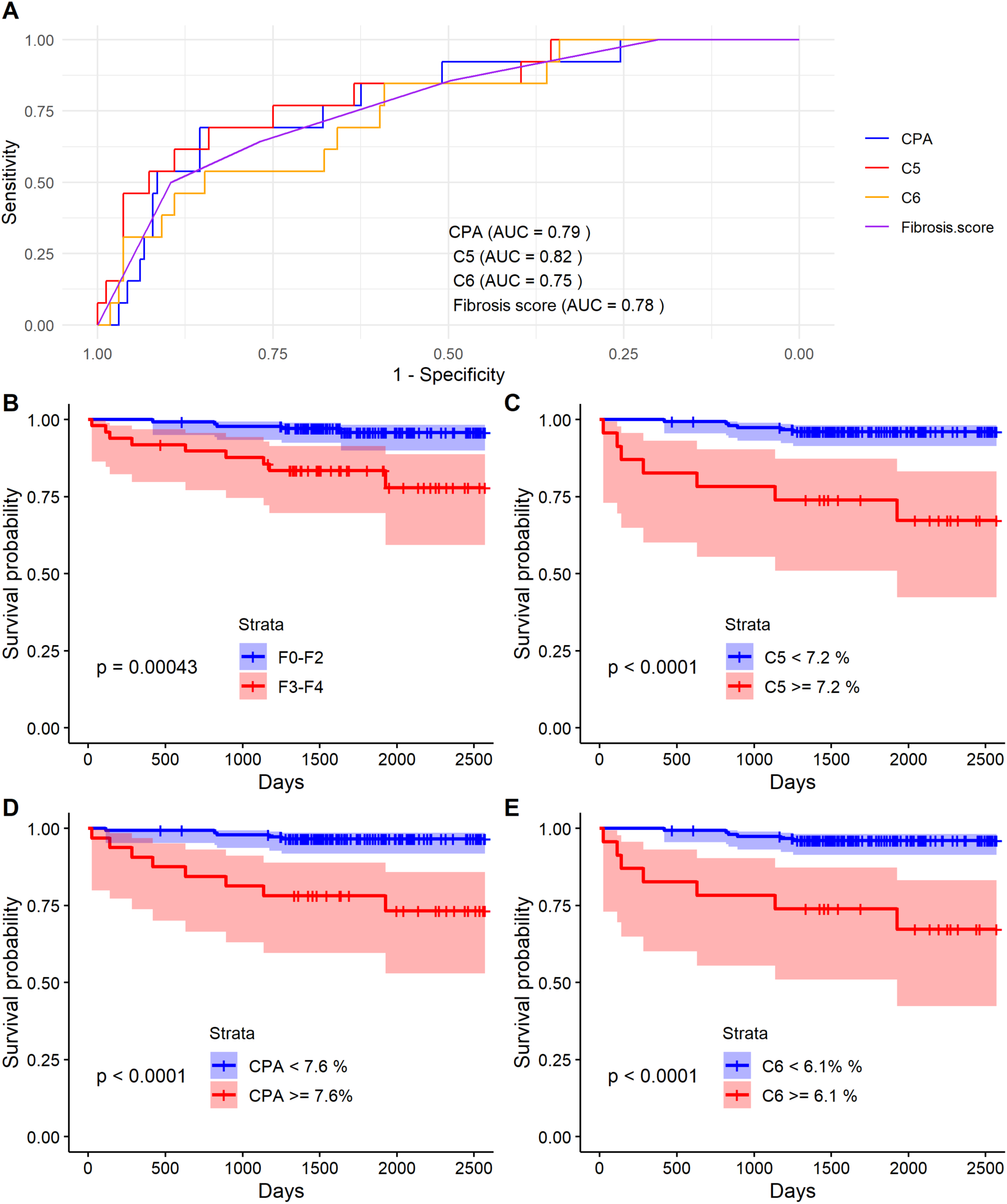
Prediction of liver-related events in the CoCoMASLD cohort using fibrosis metrics during follow-up. **A)** AUROC for prediction of liver-related events using CRN fibrosis score, CPA, and CDPs C5 and C6. **B)** Kaplan-Meier plot stratified by advanced fibrosis (F3-F4). **C)** Kaplan-Meier stratified by the Youden’s index cut off for CDP C5. **D)** Kaplan-Meier plot stratified by the Youden’s index cut off for CPA. **E)** Kaplan-Meier stratified by the Youden’s index cut off for CDP C6. P-values were calculated by log-rank test.

**Table 2.** Competing risk Cox proportional hazards models for liver-related events (LRE) during follow-up. Adjusted for age and sex. Results are shown as hazard ratios (HR) with 95% confidence intervals (CI) and p-values. Collagen clusters and CPA were stratified according to optimal thresholds identified by Youden’s Index and CRN fibrosis score by advanced fibrosis. Sensitivity and specificity were calculated based on corresponding binary cut-offs.

|  | HR (95% CI),<br>p-value | Sensitivity | Specificity |
| --- | --- | --- | --- |
| Advanced Fibrosis<br>F3-F4 vs. F0-F2 | 4.26 (1.29–14.08)<br>$p = 0.017$ | 0.64% | 0.77% |
| CPA $\geq 7.6\%$ | 6.88 (2.21–21.46)<br>$p = 0.0009$ | 69.23% | 85.45% |
| C4 $\geq 5.2\%$ | 10.66 (1.36–83.84)<br>$p = 0.024$ | 92.31 % | 65.85% |
| C5 $\geq 7.2\%$ | 10.50 (3.30–33.44)<br>$p = 0.00007$ | 69.23% | 84.15% |
| C6 $\geq 6.1\%$ | 14.05 (3.99–49.53)<br>$p = 0.00004$ | 84.62% | 59.15% |

### Predicting LSM increase at follow up

Out of 186, where both a baseline and a follow up LSM was available, 21 (11.29%) had an increase in LSM. A 1% increase in C4 and C5 were associated with increased odds ratios (OR) of having LSM increase on the latest available LSM. C4 had an OR of 1.10 (95% CI 1.01 – 1.15, p = 0.020) and C5 an OR 1.16 (95% CI 1.05 – 1.29, p = 0.005). Both CPA (OR 1.08, 95% CI 1.01 – 1.15, p = 0.016) and NASH CRN fibrosis score (OR 1.76, 1.17 – 2.64, p = 0.007) also showed significant associations with LSM increase at follow up.

## Discussion

In this study, we present CDPs, derived from AI-based analysis of histological images, as novel and biologically meaningful markers of fibrosis severity and progression in MASLD. We demonstrate that these spatially defined phenotypes, particularly CDP C5, align with established clinical endpoints, exceed traditional histological metrics such as collagen proportional area (CPA) and CRN fibrosis staging in transcriptomic and proteomic sensitivity, and may offer enhanced granularity in fibrosis assessment. Importantly, our deep learning model will be made open access to support reproducibility and broad application.

### Biological relevance of collagen deposition phenotypes

Unlike CPA, which quantifies total collagen without considering its spatial distribution or organization, and CRN fibrosis staging, which is qualitative and categorical, CDPs offer a data-driven, spatially resolved characterization of extracellular matrix (ECM) patterns based on local collagen texture and density. Although these phenotypes do not directly map to anatomical regions (e.g., portal tracts or perisinusoidal spaces), several phenotypes seem to capture features of advanced architectural remodelling, such as bridging fibrosis or dense collagen aggregates. The presence and proportion of specific CDPs therefore may reflect biologically relevant differences in fibrogenesis.

Among these, phenotype C5 emerged as a particularly robust marker of biologically advanced fibrosis. C5 showed the strongest associations with differentially expressed genes (DEGs) and circulating proteins, including markers of fibrogenic activation, ECM remodelling, and hepatic dysfunction. Regression-based transcriptomic analyses revealed that both C5 and C6 had larger effect sizes than CRN F3–F4 staging or CPA. Moreover, pathway enrichment analysis indicated that phenotype C4 and particularly C5 were enriched for ECM remodelling pathways, suggesting that these phenotypes capture dynamic fibrogenic processes.

Investigating the structure of fibrosis, in addition to its quantity, may offer a more nuanced understanding of disease mechanisms and progression. For instance, morphology patterns could indicate whether fibrosis is in a dynamic versus static state or whether certain collagen architectures respond differently to therapy. Collagen phenotypes may therefore provide complementary information not captured by CPA or CRN staging alone.

### Integration with multi-omics

Recent large-scale efforts to map the molecular landscape of MASLD have highlighted the complexity of fibrogenesis (33,34,49,50). Govaere et al. identified 31 circulating proteins associated with advanced fibrosis and steatohepatitis (34), many of which overlapped with our proteomic findings associated with phenotypes C4 and C5, including SHBG, MFAP4, SVEP1, ENPP7, GDF15, and THBS2. Among these, GDF15, MFAP4, and THBS2 have also been independently associated with fibrosis severity in MASLD by other studies (48,51–53). COLEC11, which we identified as one of the most upregulated proteins in C4, C5, and F3–F4, was likewise found to be increased in F3, and particularly in F4, fibrosis in the study by Govaere et al. (34). However, it remains largely unexplored in the context of MASLD. COLEC11 is a soluble C-type lectin previously linked to cancer cell proliferation and liver metastases (54). Interestingly, Zhang et al. found COLEC11 expression to originate from hepatic stellate cells (HSCs) in both humans and mice and demonstrated that upregulation of the closely related COLEC10 promoted collagen production in mice (55). Turning to our transcriptomic data, we observed substantial overlap with findings from other studies examining gene expression in relation to fibrosis severity in MASH. Genes such as STMN2, KRT23, FAP, FGF7, COL10A1, and SMOC2 were among those previously upregulated (56,57). Notably, STMN2, one of the most strongly upregulated genes in both C4 and C5, has been independently associated with fibrosis in several omics studies (33,49,58). Arendt et al. identified STMN2 as the gene most strongly correlated with fibrosis severity (49), while subsequent studies localized its expression to macrophages in the portal and sinusoidal regions (33). Our findings extend these observations by linking STMN2 expression to specific collagen morphology patterns identified via AI-based clustering. We also found KRT23 expression to be markedly increased across CDPs C4–C6 and in F3–F4, but not in CPA. Similarly, Kendall et al. reported a prominent increase in KRT23 expression in F4 MASLD (57). KRT23 has previously been shown to be upregulated in chronic hepatitis C(59), implicated in hepatocyte proliferation (60) and suggested as a potential therapeutic target in MASLD (44).

Importantly, while most previous studies have stratified omics data using conventional metrics such as CPA or CRN scores, our results suggest that these approaches may overlook biologically active extracellular matrix (ECM) remodelling captured by AI-based collagen phenotyping.

### Clinical implications and prognostic value

Histologically, the fibrosis stage remains the strongest predictor of clinical outcome in MASLD (9,10,61,62). Robust risk stratification is essential for guiding treatment decisions and patient management. Almost all patients who developed liver-related events (LREs) in this study had a severe disease course, with LREs primarily reflecting cirrhotic decompensation and liver-related death.

From a clinical perspective, CDPs, particularly C4 to C6, emerged as strong predictors of LREs. When stratified using optimal cut-offs, C4 to C6 showed hazard ratios above 10 for LREs. This indicates that these data-driven phenotypes capture advanced and progressive disease stages more appropriately than clusters derived from categorical grading.

Longitudinal analyses further revealed that increases in C4 and C5 were significantly associated with higher odds of clinically relevant liver stiffness progression, supporting their potential as dynamic markers of disease progression.

While C4, C5 and C6 predicted liver-related clinical events on par with CPA and CRN fibrosis staging in this cohort, their key strength was their superior sensitivity in transcriptomic and proteomic analyses. It is possible that morphology-based phenotyping may offer added discriminatory value within fibrosis stages, for example, within F4, but further studies with larger cohorts and longer follow up periods are needed to explore this potential.

### Comparison to existing fibrosis metrics

Although CRN staging and CPA remain widely used, both have notable limitations. Histopathological evaluation of liver biopsies is essential for MASH and when there is diagnostic uncertainty. However, CRN fibrosis staging, while accounting for fibrosis patterns, is qualitative and subject to significant interobserver variability (12). Moreover, it does not capture the exponential nature of fibrosis progression across stages (16) and lacks sufficient granularity (12,13), particularly within cirrhosis, where both early compensated and end-stage cirrhosis are classified similarly (63). This is clinically problematic, as median survival in early compensated cirrhosis is ∼12 years, compared to ∼2 years in decompensated cirrhosis (64). These limitations underscore the need for a more granular fibrosis metric aligned with disease biology and clinical trajectory. CPA, although quantitative and granular, lacks information on collagen organization or remodelling activity. It aggregates both fibrotic and non-fibrotic collagen, reducing biological specificity. This limitation was supported by our proteomic data, where CPA poorly identified circulating fibrosis biomarkers, whereas CDPs C4 and C5 correlated with validated markers of advanced fibrosis and steatohepatitis (e.g., MFAP4, THBS2, GDF15).

CDPs address these gaps by offering a quantitative, spatially resolved, and reproducible phenotyping approach, with increased sensitivity to molecular signatures of fibrogenesis. Unlike spatial transcriptomics, which directly maps gene expression to tissue regions, our method reflects how collagen architecture relates to the broader transcriptomic state of the tissue. This systems-level approach complements spatial omics and can be applied cost-effectively to retrospective cohorts.

### Limitations & strengths

We used liver biopsy samples acquired with 16-or 19-gauge needles, which may not fully capture the spatial heterogeneity of fibrosis across the liver. Interpretation of CDP proportions should therefore be approached with appropriate caution.

While we observed strong correlations between CDP proportions and DEGs, we cannot definitively localize gene expression changes to areas with specific collagen patterns. Unlike spatial transcriptomics, our approach does not offer cell-or structure-specific molecular localization (65). Instead, it captures how fibrotic patterns influence the overall tissue transcriptome without directly assigning gene expression to specific morphological phenotype.

Nevertheless, our modular and cost-effective design enables application in retrospective cohorts and large-scale studies. Although it does not provide cellular-level resolution, the method complements spatial transcriptomics by offering a broader systems-level perspective.

Finally, due to the cross-sectional design of our study, we could not assess how collagen phenotypes evolve or respond to interventions. Repeated biopsies are needed to validate temporal changes and clinical correlations. Further validation in larger and more diverse cohorts, including varying tissue processing protocols, is also essential to assess reproducibility, generalizability, and clinical utility.

## Conclusion

MASLD is a highly prevalent yet often underdiagnosed condition, affecting up to one in three adults, and increasingly also children (66,67). Bayesian modeling suggests that the prevalence may rise to over 55% by 2040 (68), highlighting the urgent need for effective risk stratification and accessible, yet specific, non-invasive biomarkers to identify high-risk patients while optimizing healthcare resources (4).

Our approach offers a next-generation framework for linking histological features with high-dimensional molecular data. By providing an open-source, interpretable, and spatially aware tool for fibrosis quantification, we enable a more biologically meaningful integration of tissue architecture with transcriptomic and proteomic profiles. This lays the groundwork for identifying spatially informed biomarkers with potential applications in patient stratification, monitoring of treatment responses, and future development of non-invasive diagnostics.

Our findings suggest that CDPs quantification is a precise and clinically relevant tool for fibrosis assessment in MASLD. It captures fibrosis-specific molecular signals with high sensitivity in both transcriptomic and proteomic datasets. By adding biologically and spatially informed information to established fibrosis metrics like CPA, this method has the potential to improve both our understanding and monitoring of fibrotic disease progression. Phenotyping of collagen deposition could complement existing fibrosis metrics in research and clinical settings, particularly in future studies aiming to track fibrosis evolution or identify novel biomarkers and therapeutic targets.

## Author Contributions

MW and YH developed the image analysis methodology. MW and MT jointly led the manuscript drafting and revisions. MT performed the statistical association with clinical endpoints. GM, LMH conducted the transcriptomic analysis. MW led the interpretation of imaging and omics data. MT and JMM contributed to interpretation of omics results. LLG, MT and MPW conducted participant recruitment. MV, RG, and RS provided expert level pathology assessment. LLG, EDG, DW, HH, VD, DRP, VIJ, and JR designed the study and provided feedback on the paper. CPM contributed to the study and provided feedback on the manuscript. JT, NK and LLG reviewed the manuscript and provided clinical feedback. All authors contributed to manuscript revision.

## Code Availability

Code for collagen segmentation, collagen deposition phenotype (CDP) computation, and interpretation of differential gene expression results is available at https://github.com/mkatw/decoding-fibrosis.

## Data Availability

The datasets generated and analysed during the current study that include individual patient-level data are not publicly available due to Danish legislation, which does not permit sharing with-out thorough anonymisation and prior approval. However, population-level differential gene expression results are available at https://github.com/mkatw/decoding-fibrosis. Furthermore, one anonymised biopsy image with corresponding outputs is freely available at https://doi.org/10.5281/zenodo.16967316. Additional selected anonymised data supporting the main findings are available from the corresponding author upon reasonable request.

## List of Abbreviations

CPA: collagen proportionate area
CDP: collagen deposition phenotype
MASLD: metabolic dysfunction-associated steatotic liver disease
MASH: metabolic dysfunction-associated steatohepatitis
HCC: hepatocellular carcinoma
NITs: non-invasive tests
AI: artificial intelligence
CRN: NASH Clinical Research Network

