## Supplementary material for "Decoding Fibrosis: Transcriptomic and Clinical Insights via AI-Derived Collagen Deposition Phenotypes in MASLD"

### 1. Omics discovery sub cohort population statistics

Of the 202 study participants, 130 were part of the omics discovery cohort. Notably, no healthy volunteers were part of the omics discovery sub-cohort. Figure 1 shows the distribution of age, BMI, diabetes and gender in the omics discovery group.

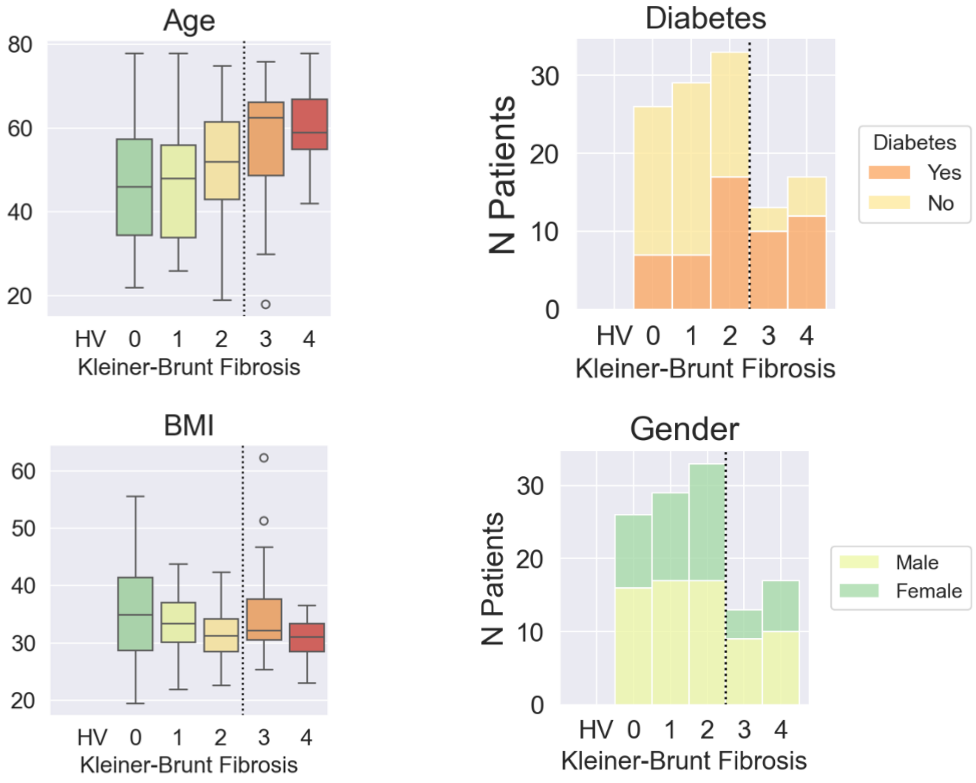

Figure 1. Population statistics of the omics discovery cohort: distributions of age, BMI, diabetes and gender by fibrosis score.

### 2. RNA sequencing and blood proteomics processing

#### Proteomics Somalogic Processing

The SomaScan v. 4.1 (7k) platform was used to generate SomaLogic data from serum samples. Relative Fluorescent units (RFU) computed by Adaptive Normalization by Maximum Likelihood (ANML) were provided by the vendor. Samples that did not pass vendor QC thresholds were removed. Likewise, aptamers classified as non-human or non-proteins or with median RFU below lower limit of detection were excluded from the analysis. RFU were log2 transformed before further analyses.

#### Bulk RNA-sequencing from FFPE samples and bioinformatics

Four 10 µm sections were cut from FFPE liver needle biopsies for RNA extraction using the RNeasy FFPE Kit (Qiagen). RNA quality was determined by measurements with the Fragment Analyzer System (Agilent, CA). Strand-specific libraries were generated from high-quality total RNA evaluated as percentage of RNA fragments longer than 200 nucleotides (DV200), higher than 30%. Coding regions of the transcriptome were captured using TruSeq RNA Exome kit (Illumina Inc. CA). Samples were subjected to single end (1 x 75bp) sequencing with NovaSeq S2 Full Flow Cell (Illumina). Reads were aligned to human reference genome (GRCh38 - hg38) using STAR 2.7.3a and read quantification was performed with Salmon 1.2.0. using Ensembl gene annotation GRCh38 (release 99). Data quality control was performed with FastQC 0.11.9, Picard CollectMultipleMetrics 2.21.6 and STAR collected and unified with multiQC. Exploratory analysis such as principal component analyses (PCA), multi-dimensional scaling and identification of confounding variables were performed after variance stabilizing transformation using DESeq2 1.26.0.

#### Bulk-RNASeq Processing and Analysis

Normalization was performed using the limma v.3.52.1 in R. The calcNormFactors function was used to calculate normalization factors for each sample. The normalization factors were then applied to the raw counts using the voom function from the limma package. The voom function performs a variance-stabilizing transformation on the data.

#### Differential Expression Analysis

Differential gene expression analysis was performed using limma v.3.52.1 R package. A linear model was fit using lmFit on normalized data while adjusting for age, sex, BMI, and diabetes status.

For transcriptomics analysis the sequencing library generation batch was included in the model to account for technical variation. The eBayes function was used to compute moderated t-statistics for continuous and categorical clinical variables. Trend and robust parameters were set to true for proteomics data (Somalogic) and set to false for transcriptomics data.

Confidence intervals were computed using the limma topTable function setting a Bonferroni-corrected threshold of 0.05. Aptamer log2 fold changes and correspondent confidence intervals were joined with the results from the mediation analysis based on SomaLogic sequence identifiers.

#### Ordinal Regression Analysis (Fibrosis Score)

Ordinal regression analysis was performed using the clm function from the R ordinal package v. 2022.11-16. The simplified fibrosis score (F0-F2 vs. F3-F4) was fit as an ordinal variable into the model, and the analysis was adjusted for library generation batch effect and other clinical variables such as age, sex, BMI, and diabetes status. The default parameters were used for the clm function. Multiple testing was accounted for by FDR adjustment.

### 3. Implementation details of the image analysis pipeline

#### Data

The imaging data consisted of 203 histology slides from 195 patients. The tissue was stained with the PicroSirius Red (PSR) staining for collagen. The glass slides were digitised at 40x magnification using Hamamatsu C13210 scanner and exported in the *.ndpi format (Figure 2). Of the 203 slides, 4 contained other stains (reticulin, others) and were rejected from analysis. Two files were corrupt. Overall 197 PSR liver histology slides were used for imaging pipeline development. Additional pre-processing (cropping) was necessary for 51 of those 197 images, as the digitised slides contained large regions of background including artefacts such as bubbles, glass slide borders, etc.

Of the 197 histology slides, 177 were used for imaging algorithm validation. 2 slides failed processing. 11 healthy volunteers did not have an accompanying fibrosis score. Further 2 patients were excluded due to incomplete or missing data. In 5 cases which had more than one PSR slice only the first slice was used for validation.

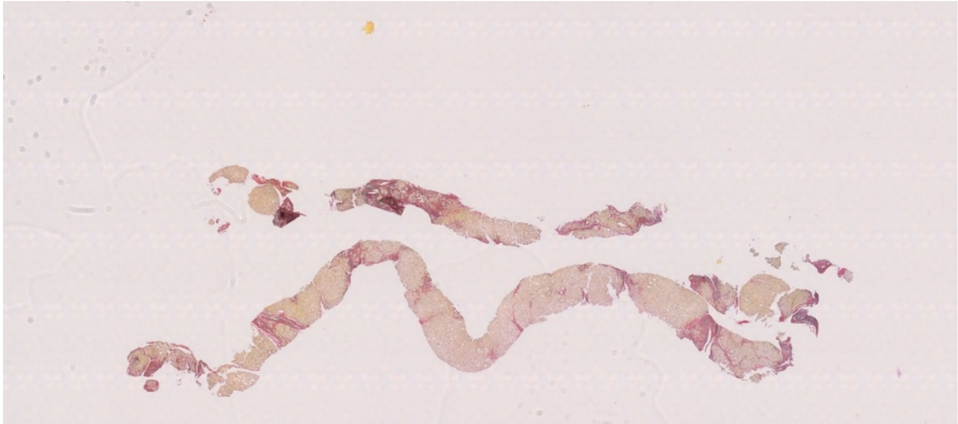

Figure 2. An example PSR-stained digital slide from the CoCoMASLD cohort.

#### Collagen segmentation

A deep-learning based approach was selected as the best solution for precise identification and segmentation of thin collagen fibres in PSR-stained slides. As chemically stained slides are often affected by stain intensity variation and colour gradients, a colour analysis of the whole PSR cohort was performed using our previously developed cohort stain characterisation tool (github.com/mkatw/slide_colour_palette). Figure 3 shows the colour distribution of all the PSR biopsies in the CoCoMASLD study. Based on the results of the colour analysis, a sub-cohort of

12 cases with maximally different staining characteristics was chosen as a representative group for collagen annotation.

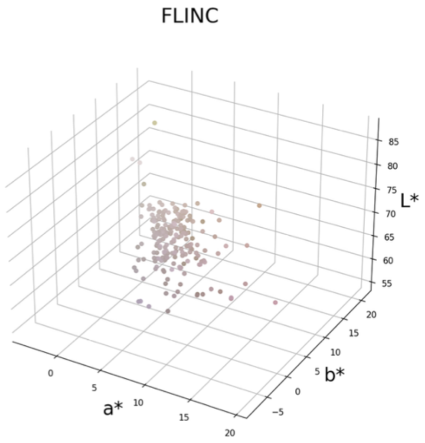

Figure 3. The distribution of CoCoMASLD PSR slide colours in the LAB colourspace. Slides both from the centre of the distribution, as well as its extremes were carefully chosen for construction of the training and testing dataset for the collagen segmentation model.

Dense annotations of collagen fibres have been performed using the approach described in our previous work[1]. Overall, 968 representative 512-by-512 px tile-annotation pairs were created. Of these, 90% randomly sampled across the annotated pool were used for algorithm training and 10% for algorithm validation. A custom version of the U-net network was implemented and trained using Python/Keras. The results of collagen segmentation for each case were saved as raw probability maps in BigTIFF format and carefully visually reviewed.

#### CPA

Collagen proportionate area (CPA) is defined as the ratio of collagen positive tissue pixels to tissue pixels. We have trained a tissue segmentation U-net analogously to the collagen segmentation model, using the same set of previously extracted tiles. Tile-level annotations were generated automatically through intensity-based thresholding. For each case used for constructing the training dataset, an Otsu intensity threshold was obtained on the case level and subsequently applied to all tiles belonging to that case.

The thus trained tissue segmentation network produced precise maps representing the probability of each pixel belonging to tissue. As histological slices are thin and include many regions of transparency (e.g. fat vesicles, blood vessel lumens), such maps required additional post-processing (Figure 4). The final tissue area was computed from “filled-in” tissue maps, in which all small holes were filled through morphological closing and filling operations.

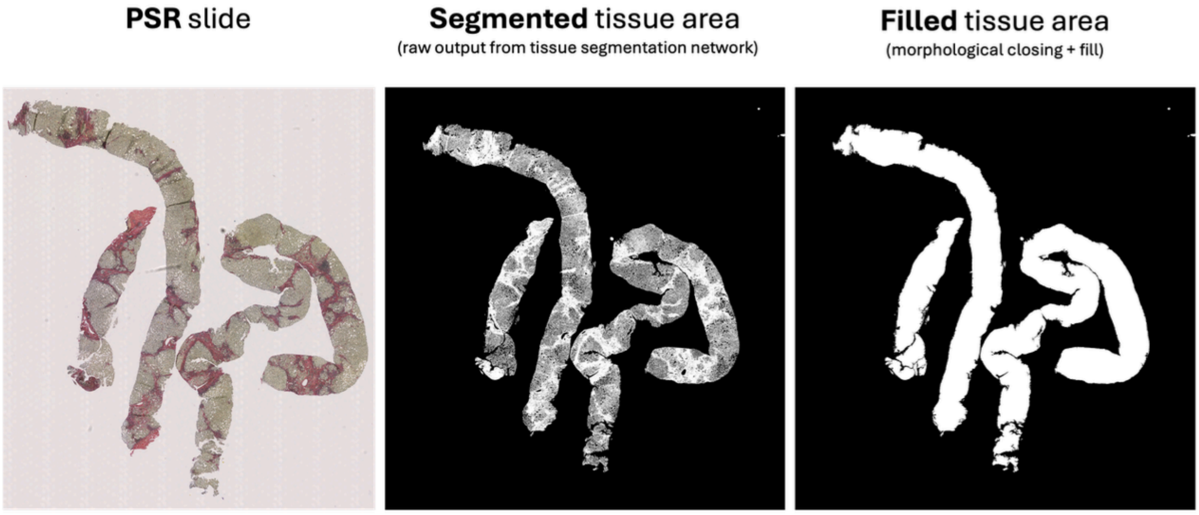

Figure 4. An example output of the tissue segmentation network, and the corresponding filled tissue map.

#### Collagen textural subtyping

To characterise the textural patterns of fibrosis in the CoCoMASLD cohort, a tile dataset has been constructed for training an unsupervised collagen tile classifier. 1 in every 10 tissue containing tiles from all the 197 PSR digital slides were extracted and pre-processed to eliminate features other than collagen (fat vesicles, cell nuclei, etc.) using the U-net network previously trained for collagen segmentation for CPA quantification. Figure 5 shows the collagen content of the extracted PSR tiles sampled randomly across the CoCoMASLD cohort.

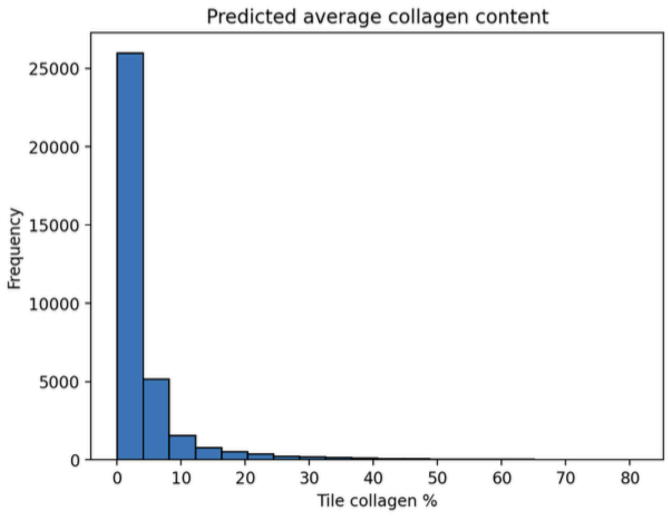

Figure 5. The distribution of collagen content in tissue tiles randomly sampled from the CoCoMASLD cohort. Most of the tiles contain very little collagen, which is consistent with pathological observation.

The extracted greyscale collagen tiles were used to learn fibrosis textural subtypes. The final randomly sampled training cohort consisted of over 3.5 thousand tiles with different collagen/fibrotic patterns. The pipeline for collagen tile clustering is summarised in Figure 6.

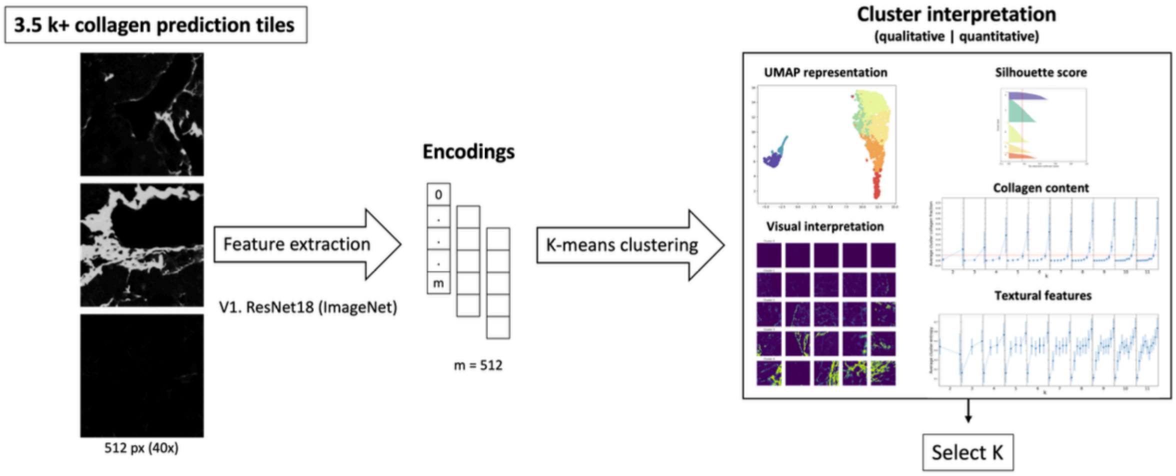

Figure 6. An overview of the image processing pipeline for unsupervised collagen pattern subtyping. Collagen predictions being the output of the trained segmentation network were used as an input for subtype learning. ResNet18 pretrained on ImageNet was used to encode the tiles into feature encodings. K-means clustering was applied to the extracted feature vectors. Qualitative and quantitative interpretation methods were applied to survey the feasible number of clusters, k.

We've applied several qualitative and quantitative methods to survey the possible number of clusters. These included mapping of the encoding feature space (PCA, UMAP) and visual interpretation of collagen tiles in each cluster. The quantitative methods included the computation of the silhouette score, average cluster collagen content and average cluster textural features (Haralick features). The clusters were always assigned numbers based on their average collagen content, with tiles in cluster 0 having the least collagen and those in cluster k the most.

The final clustering was performed iteratively, resulting in k = 5 + 2 = 7 clusters. In the first iteration, the whole cohort was divided into 5 clusters (clusters 0-4). We've found that cluster 4 (tiles with dense collagen aggregates), had a very large internal variation in collagen content. Therefore, in the second clustering iteration, we've subdivided the original cluster 4 into 3 clusters now called 4, 5 and 6, respectively. Figure 7 shows example tiles representative of each resulting cluster. The thus trained collagen tile classifier was applied to the all the PSR slides in the CoCoMASLD cohort. Figure 8 shows the mean collagen cluster profiles for the respective fibrosis stages in the study. Figure 9 shows the change in collagen cluster abundances with disease progression, relative to the mean value for healthy volunteers.

The code for the analysis as well as the trained models are available on GitHub: https://github.com/mkatw/decoding_fibrosis.

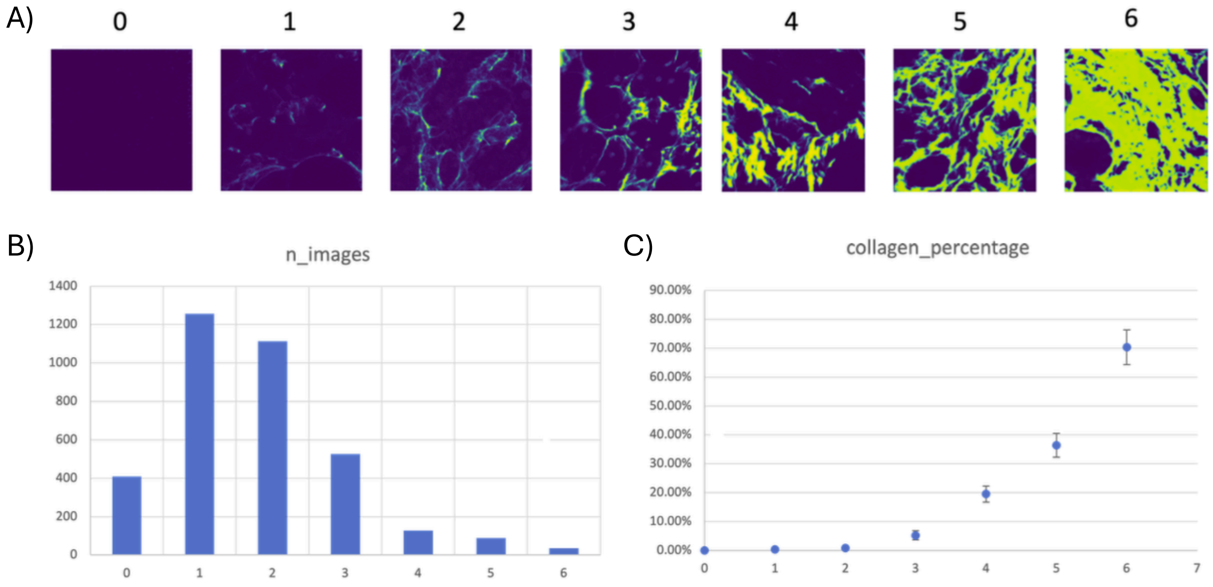

Figure 7. A) Example images representing the 7 identified collagen texture clusters. B) Distribution of number of tiles belonging to each cluster in the training cohort. C) Distribution of mean collagen percentages in tiles belonging to each cluster in the training cohort. Most tiles in the training cohort contain very little collagen (clusters 0-2). On the other hand, tiles with very densely aggregates collagen fibres (clusters 4-6) only represent a small fraction of the fibrosis stage-balanced cohort.

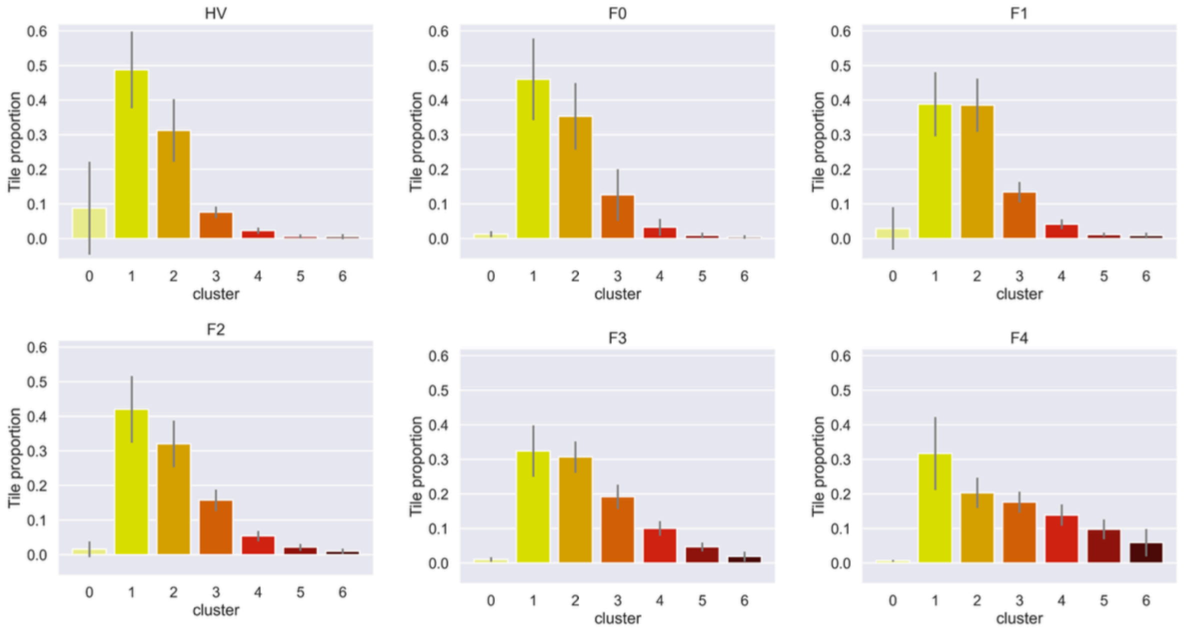

Figure 8. Mean collagen tile cluster histograms by fibrosis stage. Error bars represent standard deviation.

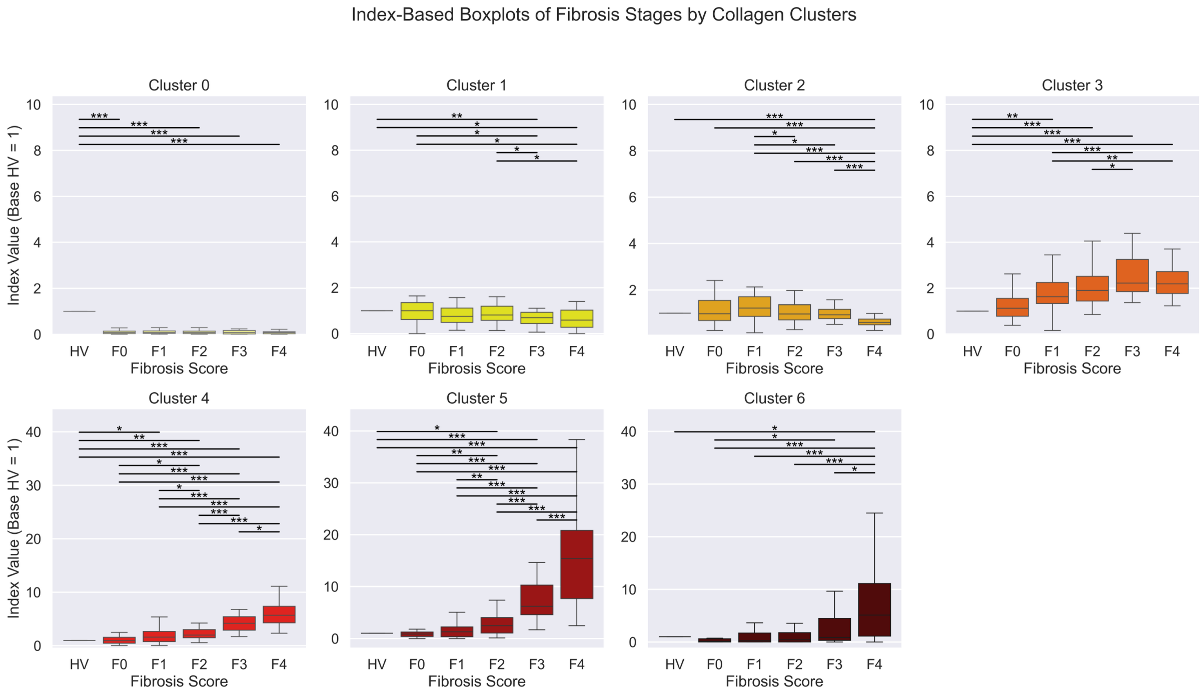

Figure 9. Mean change in cluster content relative to healthy volunteers for each collagen cluster and fibrosis score. Overall cluster 5 was found to increases the most significantly with fibrosis stage.

#### 4. Observation of Outcome

Data are presented as n (%), mean ± standard deviation (SD), or median [IQR]. All inferential tests are two-tailed with a nominal alpha level of 0.05. No adjustments for multiple comparisons were made due to the exploratory nature of the analyses. All statistical analyses were conducted using R v4.3.0, and plots were generated with the *ggplot2 (v3.5.1)* and *ggsurvfit* (v1.1.0) packages.

All participants were included in the baseline evaluation of correlations between AI-derived histology metrics, CRN fibrosis stages, and LSM. Only MASLD patients were included in the analyses of disease progression.

To test correlations between baseline fibrosis stage, LSM and continuous or ordinal histology metrics, Spearman's rank-order correlations were used. Comparison between groups were done using Wilcoxon rank-sum test.

In the time-to-event analyses, disease progression was defined as the occurrence of a liver-related event (LRE) after inclusion. LREs included liver-related death, liver transplantation, hepatocellular carcinoma, hepatic encephalopathy, hepatorenal syndrome, jaundice, ascites, variceal hemorrhage, and varices detected on endoscopy.

Using the *coxph* function from the *survival* package (v3.7-0), both univariable and multivariable competing risk Cox proportional hazards models—adjusted for age and sex—were employed to analyze the association between fibrosis metrics and LREs, considering death from causes other than cirrhosis as a competing risk.

Receiver operating characteristic (ROC) curves were generated using the pROC package (v1.18.5) to evaluate the predictive performance of fibrosis metrics for LREs. The optimal threshold for each continuous metric was determined by the Youden index. Fibrosis-related variables were dichotomized based on these thresholds, while the CRN fibrosis score was categorized as F3-F4 (advanced fibrosis) and F0-F2. The resulting cut-offs were applied to stratify the cohort, assess outcome distributions via the log-rank test, and visualize differences using Kaplan-Meier plots.

In multivariable logistic regression analyses, we used the *glm* function from the *stats* package (v4.3.0) for the binominal coded outcome of disease progression, defined as a clinically significant increase in LSM from baseline to the latest available LSM. A clinically significant increase was defined as an increase of ≥5 kPa and ≥20% from the baseline LSM value. Logistic regression models were adjusted for age and sex.

**Supplementary Table 1. Results from competing risk Cox proportional hazard analyses of LRE during follow up.** Results are expressed as hazard ratios (HR) per 1% increase for continuous variables (i.e. collagen cluster percentages and CPA), and for one level increase for categorical variables (i.e. fibrosis score).

| | Univariable | | Multivariable | |
|---|---|---|---|---|
| **Variable** | HR (95% CI) | p-value | HR (95% CI) | p-value |
| **Age** | 1.04 (1.00- 1.09) | 0.047 | | |
| **Sex (male)** | 5.02 (1.23 – 22.43) | 0.035 | | |
| **Cluster C0** | 0.87 (0.52 – 1.43) | 0.58 | 0.82 (0.45 - 1.51) | 0.53 |
| **C1** | 0.96 (0.94 – 0.99) | 0.020 | 0.96 (0.93 – 0.99) | 0.022 |
| **C2** | 0.97 (0.93 – 1.01) | 0.18 | 0.97 (0.93 – 1.01) | 0.16 |
| **C3** | 1.04 (1.00 – 1.08) | 0.026 | 1.04 (1.00 – 1.08) | 0.065 |
| **C4** | 1.15 (1.08 – 1.22) | 0.00003 | 1.12 (1.05 – 1.20) | 0.0005 |
| **C5** | 1.22 (1.13 – 1.32) | 0.000001 | 1.21 (1.11 -1.32) | 0.000001 |
| **C6** | 1.07 (1.00 – 1.15) | 0.036 | 1.13 (1.03 – 1.24) | 0.010 |
| **CPA** | 1.05 (1.01 – 1.10) | 0.020 | 1.08 (1.02 – 1.15) | 0.014 |

| | | | | |
|---|---|---|---|---|
| **Fibrosis score (F0-F4)** | 2.33 (1.46 – 3.71) | 0.0004 | 2.09 (1.26 – 3.46) | 0.004 |
| **Advanced Fibrosis (F0-F2 vs. F3-F4)** | 5.69 (1.90 – 16.98) | 0.002 | 4.26 (1.29 – 14.08) | 0.017 |
| **CPA (Youdens threshold)** | 7.84 (2.56 – 24.03) | 0.0003 | 6.88 (2.21 – 21.46) | 0.0009 |
| **C6 (Youdens threshold)** | 7.72 (2.36-25.15) | 0.0007 | 14.05 (3.99-49.53) | 0.00004 |
| **C5 (Youdens threshold)** | 8.88 (2.97 – 26.50) | 0.00009 | 10.50 (3.30 – 33.41) | 0.00007 |
| **C4 (Youdens threshold)** | 13.60 (1.77 – 104.65) | 0.012 | 10.66 (1.36 – 83.84) | 0.024 |
